# Cholesterol Paradox: Multi-point Heart Failure Prediction from Lipid Panels in Electronic Health Records

**DOI:** 10.64898/2026.10.21.26366145

**Authors:** Yina Hou, Md. Kamrozzaman Bhuiyan, Manar D. Samad

## Abstract

In cardiovascular medicine, low-density lipoprotein cholesterol (LDL-C) is the main lipid marker used in routine screening to assess cardiovascular risk and guide preventive treatment and dietary changes. An increasing number of studies have recently documented the cholesterol paradox: patients with or at risk of heart failure (HF) often have unexpectedly low levels of LDL-C despite leading to cardiovascular diseases. This paper examines how well retrospective lipid panel measurements, supplemented with apolipoprotein B (ApoB), can predict an initial heart failure (HF) event in several pre-diagnosis time windows, using age-matched patient data from Electronic Health Records (EHRs). This retrospective study uses lipid profiles from multicenter nationwide EHRs from the All of Us Research Program. We apply machine learning and explainable AI (SHAP) to lipid-based prediction and to interpret cholesterol contributions to the diagnosis of first heart failure (HF). One experiment predicts the diagnosis of HF in 36 months; the other distinguishes HF patients from people who have never been diagnosed with cardiovascular disease. A baseline lipid panel that includes ApoB predicts the first HF event up to 36 months in advance, achieving an AUC of 0.624 in men and 0.627 in women. Compared to a group of the same age without previous cardiovascular disease, a lipid profile including ApoB predicts a first HF event with a substantially higher AUC: 0.937 for men and 0.925 for women. Shapley value analyses suggest that higher triglycerides and lower HDL and LDL levels are linked to subsequent HF events, with LDL appearing as the strongest yet counterintuitive predictor. ApoB ranks second in importance; in males, higher ApoB is associated with a greater risk of HF, unlike LDL. In general, the study highlights an unexpected association with LDL and emphasizes the prognostic utility of sex-specific metabolic indicators (TG, HDL) for HF using routinely collected clinical data.

## I. Introduction

For decades, Low-Density Lipoprotein (LDL) cholesterol has been characterized as the “bad” cholesterol, and clinical care still largely relies on this indicator to assess cardiovascular disease (CVD) risk and inform preventive therapies. Medical guidelines have drawn on the association—though not necessarily a causal link—between cholesterol and cardiovascular disease, as popularized by Ancel Keys’ well-known seven-country studies in the mid-1960s [1]. Dietary saturated fats are widely accepted as a principal contributor to increased serum LDL levels, a result used to link saturated fat intake to cardiovascular disease and to support recommendations to replace them with polyunsaturated fats. As a result, current dietary guidelines generally require the limitation of saturated fatty acids without taking into account differences in biological effect, food source, structural subtype, or other molecular features. Similarly, the oversimplification of cholesterol has long misinformed medical professionals to advise against the consumption of nutrient-dense proteins such as eggs until recent confirmation in research that dietary cholesterol does not increase serum cholesterol [2].

However, cholesterol-lowering drugs, such as statins, are routinely prescribed in primary care and are known to significantly worsen markers of insulin resistance and metabolic panels [3], [4]. It should be noted that CVDs are multifactorial, where metabolic syndrome and insulin resistance play a significant role in many of the patient population. For example, the population of the Indian subcontinent has one of the highest CVD rates, despite being one of the lowest consumers of animal protein and saturated fat-rich diets. Furthermore, studies have shown that statin administration significantly reduced serum LDL levels but did not significantly improve survival rates in patients at risk of CVD [5]. Therefore, the causal role of LDL is questionable when reducing LDL alone does not provide a robust safeguard against CVD. Moreover, recent studies have revealed data supporting the ‘cholesterol paradox’ [6], [7], in which a substantial proportion of CVD patients, including heart failure patients, have normal to low levels of LDL. These results prompt a consideration of whether LDL plays a causal role in CVD, rather than simply reflecting the associative link suggested by many correlation-based studies. The cholesterol paradox reveals that cholesterol levels in the bloodstream are poorly correlated with cholesterol deposited in the blood vessel [8]. Some argue that not all cholesterol particles are equally atherogenic and that inflammation in blood vessels is often necessary for cholesterol deposition and atherosclerotic plaque formation.

Within the changing and perplexing scientific paradigms of the diet-heart and lipid hypotheses, a new marker, Apolipoprotein B (ApoB), has gained momentum and trust in the health science community for its ability to better capture CVD risk than traditional LDL concentrations. ApoB is argued to measure particle counts and is reported to more accurately assess atherosclerosis risk than standard LDL-C. Unfortunately, ApoB tests are costly and are not included in preventive care or standard screening, which are typically covered by health insurance. When ApoB measurements are unavailable at a follow-up visit, we use a robust mathematical formula from [9] to estimate ApoB levels from observed total cholesterol (TC), HDL, and triglycerides. We use baseline measurements from the standard lipid panel and ApoB to forecast the first heart failure event across different future time horizons. Experiments are conducted to elucidate and demonstrate both the predictive accuracy and the quantitative contribution of lipids to heartfailure prediction in men and women.

## II. Related work

Cardiovascular risk stratification has historically been anchored to LDL-C as the primary lipid biomarker in both clinical practice and predictive modeling [1], [5]. However, the literature suggests that LDL-C alone provides an incomplete representation of the atherogenic burden, particularly in populations with heart failure. This section reviews the relevant literature in three dimensions: documented limitations of LDL-C as a CVD risk marker, the emerging clinical and predictive utility of apolipoprotein B, and the application of explainable machine learning frameworks to assess cardiovascular risk based on lipid levels.

The fundamental challenge to LDL-C as a reliable CVD risk marker has been documented in multiple large-scale epidemiological and clinical studies. In a large American study [10], including almost 140,000 patients with CVD, their LDL-C at the time of hospital admission was actually lower than normal. Gomes et al. [11] summarized multiparametric models for the stratification of risk in patients with HF, suggesting that LDL-C is not a predictor of the severity or progression of the HF disease. In a cohort of 1,274 patients with chronic stable HFrEF (Heart Failure with reduced ejection fraction) from the study [12], the overall median LDL-C was only 89.0 mg/dL, with 60.7% of patients falling below the 100 mg / dL threshold, underscoring the well-documented tendency for HF patients to present paradoxically low levels of LDL-C despite elevated cardiovascular risk.

Paradoxically, higher LDL-C has been associated with better survival in chronic and acute HF [13], [14], a finding that contradicts the foundational assumptions of lipid-lowering therapy. Statin administration, while effective in reducing LDL-C, has not demonstrated robust improvements in allcause survival for broad at-risk populations [5], further eroding confidence in LDL-C as a causal risk marker rather than a correlate. Together, these findings motivate the investigation of alternative lipid biomarkers with a stronger mechanistic and prognostic grounding. Recent studies have explored lipid biomarkers beyond LDL-C to improve cardiovascular risk assessment. In particular, apolipoprotein B (ApoB), which reflects the total number of circulating lipoprotein particles, has been reported to provide stronger associations with cardiovascular outcomes than LDL concentration alone. Wong et al. [15] integrated ApoB into the Systematic Coronary Risk Evaluation 2 (SCORE2) cardiovascular risk framework using 448,303 UK Biobank participants, finding that while ApoB alone showed limited standalone discriminatory performance, it demonstrated a complementary role beyond conventional lipid indices. A comprehensive review [16] shows that ApoB better predicts atherosclerotic cardiovascular disease risk than LDL-C by directly quantifying atherogenic lipoprotein particle number, with superior precision in patients with discordant lipid profiles, hypertriglyceridemia, or very low LDL-C. Sexspecific differences in lipoprotein metabolism further suggest that ApoB may have a differential prognostic value between male and female populations [16], a dimension that has remained underexplored in previous studies of HF prediction. Despite its demonstrated superiority, ApoB measurement remains largely absent from routine clinical screening due to cost and limited insurance coverage. Hwang et al. [9] derived a validated mathematical formula to estimate ApoB concentration in mg/dL directly from a standard lipid panel, including TC, HDL-C, and triglycerides, enabling its use in large-scale EHR datasets where direct ApoB measurements are rarely available.

Although prior studies have established the prognostic relevance of LDL-C and ApoB using traditional survival methods such as Cox regression and Kaplan-Meier analysis, these approaches share key limitations: they rely on binary lipid thresholds, model one biomarker in isolation, assume linear associations, and evaluate only a single time point relative to diagnosis. Explainability has become a central concern in clinical machine learning, as predictive accuracy alone is not sufficient to support clinical adoption. The SHapley Additive exPlanations (SHAP) framework [17] provides theoretically principled feature attribution scores for tree-based models, allowing clinically interpretable explanations of individual predictions. SHAP-based analysis has been applied to identify biomarker contributions in cardiovascular risk models [18], but its use for lipid biomarkers in multiple pre-diagnosis time windows in HF, with sex-stratified analysis, has not been explored. This paper addresses these gaps by applying XGBoost with SHAP attribution to characterize the nonlinear, multi-biomarker, sex-specific, and temporally varying signal within the lipid panel for HF prediction using large-scale EHR data.

The contributions of this paper are as follows. First, this paper presents one of the first applications of explainable machine learning to predict an initial HF diagnosis using retrospective lipid panel data. Second, we integrate ApoB, a more recently identified predictor of cardiovascular disease risk, into the prediction model to clarify its role in predicting HF. Third, we evaluate multiple models to predict future HF diagnoses in different time windows to understand the effect of time. Fourth, we compare people who later develop HF, observed at different intervals before their initial diagnosis, with people without cardiovascular disease to identify the lipid markers that distinguish these two cohorts.

## III. Methodology

This section describes the human subject data, IRB exemption for secondary use of de-identified human subject data, the electronic health record curation process, the ApoB estimation formula, the patient labeling strategy for two predictive tasks, and the evaluation protocol. All methods were performed in accordance with the relevant guidelines and regulations, as elaborated below.

### A. Human subject data and ethics statement

This study used secondary, de-identified human subject data available through the credentialed repository of the All of Us Research Program in the United States [19]. The All of Us program collected these data exclusively for research, and participants provided their informed consent at enrollment. Because the data used here are de-identified, the protocol governing secondary use does not require additional consent. The use of human subject data was reviewed by the Institutional Review Board (IRB) at North Carolina Agricultural and Technical State University (IRB protocol number HS26-0215) and was found to be exempt from additional review under the regulatory category in 45 CFR 46.101(b). The IRB granted this exemption based on the Category 4 criteria. The study will involve Exempt (4ii): (4) Secondary research for which consent is not required: Secondary research uses of identifiable private information or identifiable biospecimens, if at least one of the following criteria is met: 1)The identifiable private information or identifiable biospecimens are publicly available, and 2) Information, which may include information about biospecimens, is recorded by the investigator in such a manner that the identity of the human subjects cannot readily be ascertained directly or through identifiers linked to the subjects, the investigator does not contact the subjects, and the investigator will not re-identify subjects.

### B. Curation of EHR data

Structured EHR data are obtained from the All of Us Research Program, a nationally representative longitudinal cohort that gathers electronic health records, survey data, and physical measurements from enrolled participants throughout the United States [19]. Patients are identified in two cohorts: 1) patients with a recorded heart failure (HF) diagnosis at any point in time and 2) patients without any record of cardiovascular disease (non-CVD), including HF. Patients with an HF diagnosis are identified using the set of ICD-10 codes under category I50, listed in Table I. The contrasting non-CVD cohort is obtained by selecting All of Us patients with recorded ICD-10 diagnoses and excluding those with a heart-related cardiovascular condition. These conditions are a subset of the I00–I99 chapter for diseases of the circulatory system [20]. Four lipid biomarkers are extracted for each patient record: low-density lipoprotein cholesterol (LDL-C), highdensity lipoprotein cholesterol (HDL-C), triglycerides (TG), and estimated apolipoprotein B (ApoB). We also retrieve total cholesterol (TC) from the EHR data and use it only to estimate ApoB and impute missing LDL-C values. Most patient records do not include the ApoB measurement. Therefore, ApoB is estimated using the formula derived by Hwang et al. [9] following Equation 1.

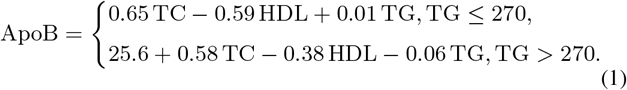

**TABLE I.** List of International Classification of Diseases, 10th Revision (ICD-10) codes corresponding to different heart failure (HF) diagnoses used in this study.

| Diagnosis | ICD-10 Codes |
| --- | --- |
| General HF | I50 |
| Systolic (congestive) HF | I50.2–I50.23 |
| Diastolic (congestive) HF | I50.3–I50.33 |
| Combined systolic and diastolic HF | I50.4–I50.43 |
| Right HF (including due to left HF) | I50.81–I50.814 |
| Biventricular / High output / End stage HF | I50.82–I50.84 |
| Other specified HF | I50.8, I50.89 |
| Unspecified HF | I50.9 |

Here, TC, HDL, and TG stand for total cholesterol, highdensity lipoprotein, and triglycerides, respectively. All values are expressed in mg/dL. We developed ApoB regression equations using multiple regression on data from individuals who underwent routine health examinations at the Samsung Kangbuk Hospital Health Screening Center between January and December 2009 and subsequently validated them in the NHANES 2007-2008 subgroups, including participants with diabetes and atherogenic dyslipidemia.

### C. Multi-time point patient records

Six pre-diagnosis windows (pre-DW) are defined at 6, 12, 18, 24, 30, and 36 months prior to the first HF diagnosis to examine how the lipid profile changes leading to the first HF event. Table II and Table III present the cohort sizes and the distributions of lipid biomarkers with mean and standard deviations for different time windows. The mean age of patients with HF in the different time windows varies from 62.79 (12.88) to 63.64 (12.21) years for males and from 61.05 (13.16) to 61.89 (12.58) years for females. To ensure age comparability between the two groups, the non-CVD cohort is iteratively selected until the difference in mean age relative to the HF cohort is no longer statistically significant (*p >* 0.05). The mean age in the non-CVD cohort is 60.84 (7.61) to 61.84 (7.17) for females and 62.68 (7.37) to 63.66 (6.91) for males. Because this age matching is performed separately against the HF cohort of each pre-DW, the matching procedure includes several non-CVD subsets for pre-DWs.

**TABLE II.**
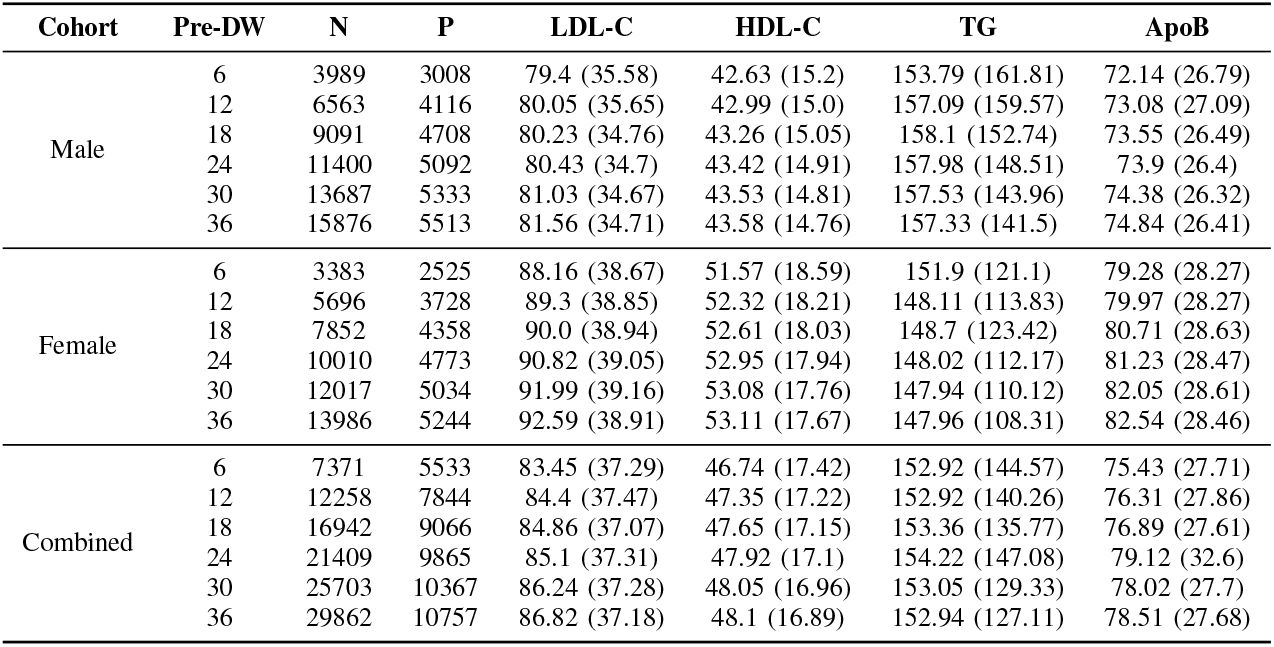
Summary of lipid panel extracted from the All of Us HF patientsstratified by sex andpre-diagnosiswindow (months) before first HF diagnosis. N = number of records. P = numberof patients.

**TABLE III.** Summary of lipid panel extracted from the All of Us non-CVD patients stratified by sex. N = number of records. P = number of patients.

| Cohort | Pre-DW | N | P | LDL-C | HDL-C | TG | ApoB |
| --- | --- | --- | --- | --- | --- | --- | --- |
| Male | 6 | 98893 | 14418 | 104.61 (33.37) | 48.86 (14.15) | 137.50 (96.09) | 87.08 (23.60) |
|  | 12, 18 | 94805 | 13886 | 104.21 (33.28) | 48.94 (14.17) | 136.76 (94.72) | 86.79 (23.51) |
|  | 24-36 | 90562 | 13333 | 103.71 (33.22) | 49.01 (14.17) | 135.98 (92.62) | 86.49 (23.42) |
| Female | 6 | 175550 | 28620 | 114.74 (34.56) | 61.08 (17.81) | 127.34 (81.39) | 94.47 (24.38) |
|  | 12 | 168259 | 27582 | 114.59 (34.57) | 61.21 (17.84) | 127.23 (80.95) | 94.42 (24.37) |
|  | 18-36 | 160702 | 26509 | 114.42 (34.54) | 61.36 (17.87) | 126.99 (80.03) | 94.34 (24.34) |
| Combined | 6 | 271107 | 42518 | 108.83 (34.46) | 56.59 (17.56) | 130.68 (86.65) | 91.58 (24.12) |
|  | 12-36 | 247823 | 39298 | 108.34 (34.43) | 56.79 (17.62) | 129.89 (84.34) | 91.23 (24.07) |

The cohort of patients with HF with a confirmed diagnosis expands from *N* = 7,371 using a 6-month lookback window to *N* = 29,862 with a 36-month lookback, due to the increased availability of laboratory data over longer time periods. By comparison, the age-matched non-CVD cohort contains 247,823 and 271,107 records across the pre-DWs. Compared to patients with HF, patients without CVD have substantially higher levels of LDL-C (108–109 vs. 83–87 mg/dL) and HDL-C (56–57 vs. 47-48 mg / dL), as well as higher estimated ApoB concentrations (91–92 vs. 75-79 mg / dL), in line with the so-called cholesterol paradox. Individuals who continue to develop HF show lower, rather than elevated, circulating lipid levels during the period preceding diagnosis.

### D. Experimental design

EHR data are curated to support two predictive tasks aligned with two research questions, as illustrated in Figure 1. The first task examines whether lipid biomarkers exhibit a timevarying predictive signal in individuals at risk for HF. For an N-month HF prediction horizon, where N ∈ {6, 12, 18, 24, 30, 36}, positive examples consist of lipid measurements collected within N months of the first HF diagnosis. All lipid measurements obtained earlier than N months before diagnosis are categorized as negative examples. The second task predicts the first diagnosis of HF within various time windows against patients who do not have CVD recorded at any time. Accordingly, lipid profiles obtained within N months before the first diagnosis of HF are designated as positive samples, and these are contrasted with lipid profiles from patients without any documented CVD, which serve as negative samples.

**Fig. 1.**
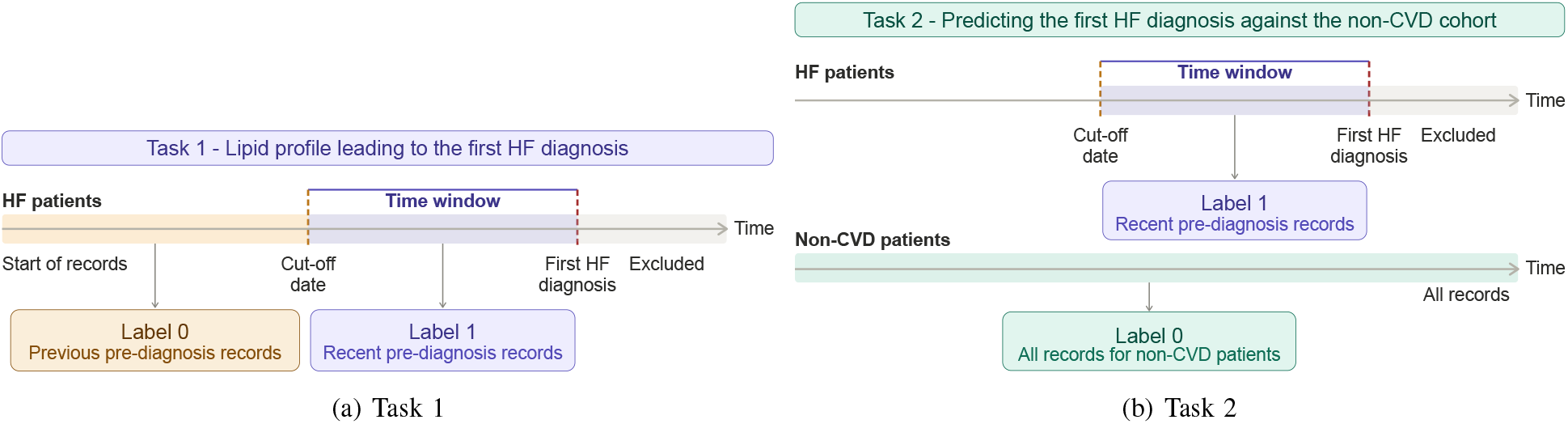
Overview of the dataset construction process for two tasks.

### E. Missing Value Imputation

Missing values are first handled using a three-stage imputation pipeline. In the first stage, a missing LDL-C value in a patient record is obtained using the Friedewald equation [21] below, when the corresponding TC, HDL, and TG values are

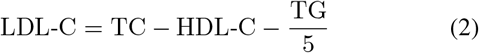

This equation is applied only to records with *TG* ≤400*mg/dL*, consistent with the validated range of the formula. This step reduces the overall missing rate from 14.52% to 9.12% in the heart failure cohort and from 12.86% to 6.38% in the non-CVD cohort. In the second stage, the remaining missing values are imputed using the median values within individual patients. Following this within-patient imputation, only 1.88% and 1.87% of lipid measurements remain missing for the HF and non-CVD cohorts, respectively. The third stage to complete the dataset is described in Section III-G.

### F. Prediction and explanation

XGBoost is used as the classifier model in all predictive tasks due to its superiority in tabular data and its robustness against outliers, overfitting, sparse features, and class imbalance compared to other models [22]. A trained XGBoost model maps each lipid record *x* to a log-odds prediction *f* (*x*). The class imbalance in data is handled using the `scale_pos_weight`parameter of the XGBoost model. To attribute this prediction to an individual biomarker *j*, we apply the SHAP framework [17], which decomposes the prediction *f* (*x*) into additive feature contributions based on Shapley values *ϕ*_*j*_ as in Equation 3.

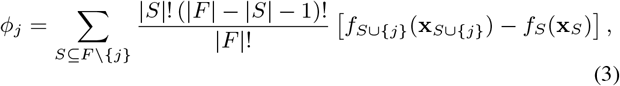

Evaluating Equation 3 exactly requires summing over all feature subsets, which is exponential in the number of features. Since XGBoost is an ensemble of decision trees, we use TreeSHAP [23] to compute the exact Shapley values in polynomial time by exploiting the tree structure, making the attribution tractable for the trained model. Shapley values can be visualized in beeswarm plots to show how altering feature values influences prediction results. In these plots, each dot represents the SHAP value of a feature for a single sample; its horizontal position indicates the signed impact on the prediction, while its color indicates the feature value from low to high. This makes it clear whether increased or decreased levels of a particular lipid push the prediction in a specific direction.

### G. Evaluation protocol

Patient samples drawn from the EHR are processed using a specific approach to address class imbalance and enhance the predictive model’s ability to generalize. In the available data, records from patients without cardiovascular disease substantially exceed those of patients with at least one HF diagnosis (271,107 combined non-CVD records versus 7,371 HF records; see Tables II and III). To train the XGBoost classifier, the non-CVD cohort is resampled 20 times using different random seeds, and in each resample the number of non-CVD patients is set to match the number of HF patients. Although this balances the cohorts by patient count, the total number of records per cohort may still differ, potentially leaving residual class imbalance in the dataset.

The classification model is assessed using a 10 × 2 nested cross-validation framework. In the outer loop, the dataset is divided into 10 folds; each fold is sequentially held out as the test set for performance assessment, while the other nine folds are used for model training. Because each patient contributes multiple follow-up records, observations are grouped at the patient level, and a patient-wise stratified k-fold split is applied so that records from the same patient never appear in different folds. This approach mitigates leakage of patientspecific information between training and test partitions. The missing values remaining after the first two imputation steps (Section III-E) are subsequently addressed within the training and test splits. Specifically, the median of each lipid variable is computed using only the training data and is then used to impute missing values in both the training and test folds.

Within each training partition, a two-fold inner validation loop is run to select hyperparameters. In the validation step, the XGBoost classifier is tuned by a grid search over a set of model hyperparameters, including the number of boosting rounds (`n_estimators` ∈ {200, 400, 500}), tree depth (`max_depth` ∈ {3, 5, 6}), learning rate (`learning_rate` ∈ {0.05, 0.1}), row subsampling (`subsample` ∈ {0.8, 1.0}), column subsampling per tree (`colsample_bytree` ∈ {0.8, 1.0}), and minimum child weight (`min_child_weight` ∈ {5, 10, 20}). Area under the receiver operating characteristic curve (AUROC) is used to select the best hyperparameter setting in the validation step. The hyperparameter configuration with the best mean inner-fold AUROC is refit using the full outer-training set and then assessed on the associated outer-test fold. Overall classification performance is reported by averaging the scores over ten outer-test folds. Along with AUROC, accuracy, F1 score, precision, recall, sensitivity, and specificity are used to characterize performance. For the explainability analysis, we calculate SHAP values using only predictions from the outer test sets, so the attributions reflect generalization performance rather than in-sample fitting. Within each cohort, we aggregate the per-instance SHAP values across all 10 outer folds and 20 random seeds into a single beeswarm summary, ensuring that the presented feature attributions are based solely on out-of-sample data.

## IV. Results

All experiments using All of Us research data must be conducted within the program’s secure cloud-based Researcher Workbench to protect the privacy of All of Us participants. We provision a cloud computing environment with a 2-core CPU with 7.5 GB of RAM within the designated workbench.

### A. Validation of estimated ApoB

Apolipoprotein B testing is relatively rare in the All of Us program, with ApoB levels measured in only 905 participants (1632 records). In contrast, estimated ApoB values are measured using the lipid panel drawn on the same day using Equation 1. In Figure 2, we validate the formula-based estimation of ApoB against its ground truth values. The estimated ApoB values show close agreement with the laboratory values measured directly. The two measures are highly correlated (Pearson *r* = 0.891, *p <* 0.001), with a small average bias of −2.27 mg/dL. The mean absolute error is 8.70 mg/dL, corresponding to a mean absolute percentage error of 9.52%.

**Fig. 2.**
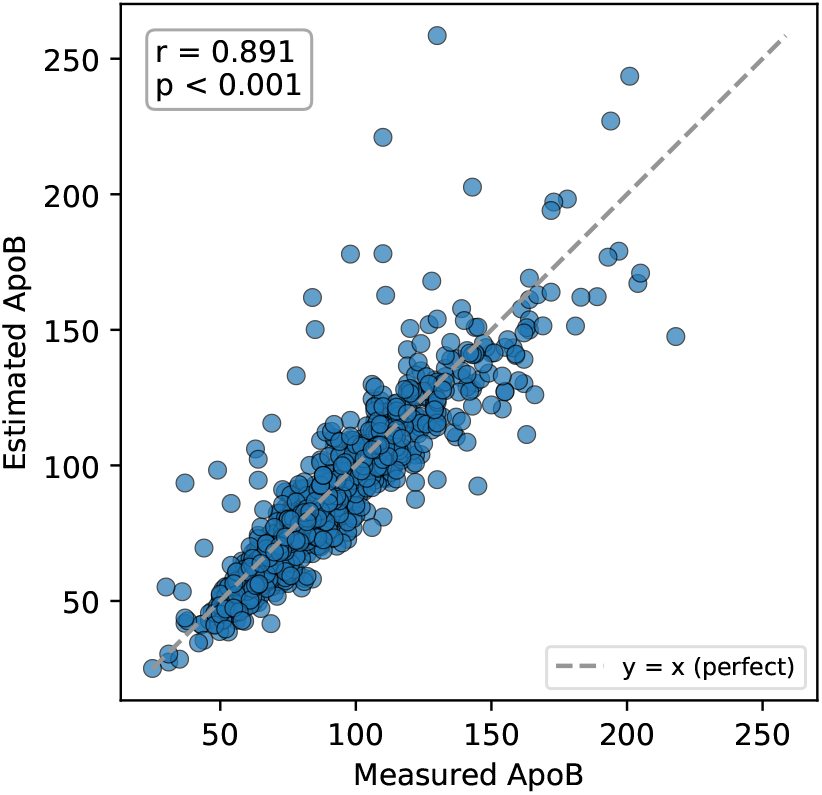
Correlation between model estimated and actual ApoB measurements.

### B. Predicting performance of lipid profiles

1. *Task I: Lipid profile leading to the first HF diagnosis:* Table IV reports the performance metrics of the XGBoost model for task I, in which the first HF diagnosis is predicted using lipid panels from six time windows preceding the diagnosis. In the combined cohort, AUROC values decline as the prediction window increases, falling from 0.677 at 6 months to 0.627 at 36 months. Accuracy (from 0.691 to 0.602) and specificity (from 0.719 to 0.627) follow a similar downward trend. Taken together, these three metrics show that the model is most accurate when the prediction window is closest to the time of diagnosis, and that the distinctive lipid signal diminishes in records that are further removed from the event. As the window widens, the records collected before the diagnosis date are absorbed into the positive class. Recall is the exception, staying nearly constant across windows (0.517–0.557), indicating the model recovers a stable fraction of positive records regardless of window boundaries. In contrast, precision and F1 increase with window size, from 0.293 to 0.430 and from 0.383 to 0.483, respectively. One possible explanation for the rise in precision, and consequently in F1 score, is that longer windows include a greater proportion of positive records. As the window length increases, each patient contributes more positive instances, which is reflected in the growth of the positive class from 7,371 records at 6 months to 29,862 records at 36 months. The male and female cohorts exhibit the same patterns in performance metrics: AUROC, accuracy, and specificity decrease as the prediction window grows, while precision and F1 increase. All performance scores comparing any pair of time windows are statistically significant across cohorts, with one exception: the AUROC scores for the female cohort between the 6-month and 36-month windows do not differ significantly (Wilcoxon signed-rank test, *p >* 0.05).
2. *Task II: Predicting the first HF diagnosis against the non-CVD cohort:* Table V reports the prediction scores for Task 2, which distinguishes patients with HF records across different temporal windows from patients who have never been diagnosed with CVD. As in Task 1, the AUROC decreases from 0.762 at 6 months to 0.728 at 36 months. Accuracy and specificity likewise decline as the prediction window lengthens. Recall stays nearly constant across windows (0.650–0.655), reproducing the pattern observed in task I. Predictive models trained within sex-specific cohorts show markedly superior performance (female mean AUROC 0.919–0.925, male mean AUROC 0.930-0.937) compared with models trained on the combined cohort (mean AUROC 0.762–0.728), aligning with the well-established differences in lipid distributions between male and female patients [24]. Unlike the combined cohort, the sex-specific models do not deteriorate as the prediction window lengthens, with the AU-ROC improving marginally from 0.930 to 0.937 in males and from 0.919 to 0.925 in females. The mean accuracy follows the same upward trend, rising from 0.821 to 0.848 in males and from 0.806 to 0.826 in females. Only in the earliest (6 months) and latest (36 months) pre-diagnosis windows do AUROC scores differ significantly between male and female patients (Mann–Whitney U test, *p <* 0.001).

**TABLE IV.** Mean area under the ROC (AUROC) curve, accuracy (ACC), F1, precision, recall, and specificity scores for prediction task I. Pre-diagnosis window (Pre-DW) is in months. Values in parentheses denote standard deviation.

| Cohort | Pre-DW | AUROC | Accuracy | F1 | Precision | Recall | Specificity |
| --- | --- | --- | --- | --- | --- | --- | --- |
| Male | 6 | 0.649 (0.017) | 0.657 (0.016) | 0.342 (0.023) | 0.250 (0.022) | 0.549 (0.016) | 0.677 (0.018) |
|  | 12 | 0.632 (0.013) | 0.635 (0.016) | 0.376 (0.014) | 0.288 (0.017) | 0.544 (0.019) | 0.659 (0.021) |
|  | 18 | 0.621 (0.010) | 0.613 (0.016) | 0.397 (0.013) | 0.314 (0.016) | 0.542 (0.022) | 0.636 (0.025) |
|  | 24 | 0.622 (0.007) | 0.611 (0.011) | 0.425 (0.013) | 0.354 (0.014) | 0.534 (0.025) | 0.640 (0.021) |
|  | 30 | 0.621 (0.008) | 0.601 (0.013) | 0.448 (0.010) | 0.386 (0.012) | 0.538 (0.021) | 0.628 (0.026) |
|  | 36 | 0.624 (0.007) | 0.595 (0.009) | 0.474 (0.011) | 0.417 (0.010) | 0.552 (0.027) | 0.616 (0.025) |
| Female | 6 | 0.682 (0.015) | 0.700 (0.016) | 0.387 (0.016) | 0.306 (0.017) | 0.534 (0.026) | 0.735 (0.021) |
|  | 12 | 0.655 (0.008) | 0.671 (0.013) | 0.402 (0.011) | 0.330 (0.013) | 0.517 (0.022) | 0.712 (0.020) |
|  | 18 | 0.644 (0.012) | 0.648 (0.012) | 0.417 (0.014) | 0.353 (0.016) | 0.514 (0.023) | 0.692 (0.019) |
|  | 24 | 0.644 (0.009) | 0.638 (0.009) | 0.447 (0.013) | 0.396 (0.015) | 0.515 (0.015) | 0.687 (0.013) |
|  | 30 | 0.637 (0.006) | 0.617 (0.010) | 0.468 (0.009) | 0.417 (0.009) | 0.535 (0.021) | 0.654 (0.021) |
|  | 36 | 0.627 (0.009) | 0.607 (0.010) | 0.479 (0.015) | 0.440 (0.013) | 0.527 (0.023) | 0.649 (0.020) |
| Combined | 6 | 0.677 (0.011) | 0.691 (0.010) | 0.383 (0.016) | 0.293 (0.015) | 0.557 (0.017) | 0.719 (0.012) |
|  | 12 | 0.647 (0.007) | 0.664 (0.011) | 0.390 (0.010) | 0.311 (0.012) | 0.526 (0.014) | 0.699 (0.017) |
|  | 18 | 0.634 (0.010) | 0.641 (0.012) | 0.408 (0.009) | 0.338 (0.011) | 0.517 (0.016) | 0.680 (0.018) |
|  | 24 | 0.634 (0.006) | 0.629 (0.012) | 0.436 (0.007) | 0.374 (0.011) | 0.524 (0.018) | 0.669 (0.021) |
|  | 30 | 0.631 (0.006) | 0.612 (0.010) | 0.462 (0.007) | 0.402 (0.010) | 0.543 (0.019) | 0.643 (0.021) |
|  | 36 | 0.627 (0.005) | 0.602 (0.007) | 0.483 (0.006) | 0.430 (0.008) | 0.553 (0.017) | 0.627 (0.017) |

**TABLE V.** Mean area under the ROC(AUROC) curve, accuracy (ACC), F1, precision, recall, and specificity scores for prediction task II. Pre-diagnosis window (Pre-DW) is in months. Values in parentheses denote standard deviation.

| Cohort | Pre-DW | AUROC | Accuracy | F1 | Precision | Recall | Specificity |
| --- | --- | --- | --- | --- | --- | --- | --- |
| Male | 6 | 0.930 (0.003) | 0.821 (0.007) | 0.619 (0.010) | 0.475 (0.013) | 0.891 (0.010) | 0.807 (0.009) |
|  | 12 | 0.932 (0.002) | 0.831 (0.005) | 0.664 (0.007) | 0.533 (0.009) | 0.883 (0.006) | 0.819 (0.006) |
|  | 18 | 0.933 (0.002) | 0.840 (0.003) | 0.707 (0.005) | 0.592 (0.006) | 0.879 (0.006) | 0.829 (0.005) |
|  | 24 | 0.935 (0.002) | 0.845 (0.004) | 0.738 (0.006) | 0.636 (0.008) | 0.880 (0.006) | 0.833 (0.005) |
|  | 30 | 0.936 (0.002) | 0.846 (0.004) | 0.759 (0.006) | 0.666 (0.007) | 0.885 (0.006) | 0.832 (0.004) |
|  | 36 | 0.937 (0.002) | 0.848 (0.003) | 0.776 (0.005) | 0.691 (0.006) | 0.887 (0.004) | 0.831 (0.004) |
| Female | 6 | 0.919 (0.004) | 0.806 (0.008) | 0.627 (0.013) | 0.480 (0.014) | 0.905 (0.008) | 0.785 (0.009) |
|  | 12 | 0.922 (0.003) | 0.808 (0.005) | 0.657 (0.008) | 0.513 (0.009) | 0.914 (0.006) | 0.782 (0.006) |
|  | 18 | 0.923 (0.003) | 0.814 (0.006) | 0.693 (0.008) | 0.560 (0.010) | 0.911 (0.005) | 0.786 (0.008) |
|  | 24 | 0.925 (0.003) | 0.821 (0.006) | 0.723 (0.007) | 0.603 (0.009) | 0.904 (0.005) | 0.793 (0.008) |
|  | 30 | 0.925 (0.003) | 0.824 (0.005) | 0.744 (0.007) | 0.633 (0.008) | 0.904 (0.004) | 0.793 (0.006) |
|  | 36 | 0.925 (0.003) | 0.826 (0.005) | 0.761 (0.006) | 0.656 (0.008) | 0.906 (0.005) | 0.790 (0.006) |
| Combined | 6 | 0.762 (0.003) | 0.722 (0.005) | 0.450 (0.005) | 0.343 (0.007) | 0.655 (0.007) | 0.736 (0.007) |
|  | 12 | 0.748 (0.004) | 0.703 (0.004) | 0.465 (0.005) | 0.363 (0.005) | 0.650 (0.006) | 0.716 (0.006) |
|  | 18 | 0.741 (0.004) | 0.692 (0.004) | 0.492 (0.005) | 0.396 (0.005) | 0.651 (0.006) | 0.705 (0.005) |
|  | 24 | 0.737 (0.004) | 0.686 (0.003) | 0.514 (0.004) | 0.426 (0.005) | 0.650 (0.006) | 0.698 (0.005) |
|  | 30 | 0.732 (0.004) | 0.678 (0.004) | 0.534 (0.005) | 0.452 (0.005) | 0.651 (0.008) | 0.689 (0.006) |
|  | 36 | 0.728 (0.003) | 0.672 (0.003) | 0.550 (0.004) | 0.475 (0.005) | 0.654 (0.007) | 0.680 (0.006) |

Overall, prediction scores for male patients (0.930–0.937) are higher than those for female patients (0.919–0.925). The consistently higher predictive scores observed in male patients compared to female patients across all time windows suggest that lipid profiles may possess stronger predictive utility in males. In the female cohort, recall consistently exceeds that of males in all time windows (0.905 vs. 0.891 at 6 months, to 0.906 vs. 0.887 at 36 months), while specificity is consistently lower (0.785 vs. 0.807 at 6 months). Thus, the model captures a greater number of true HF cases in females at the cost of more false positives.

### C. Interpretation of XGBoost and SHAP

1. *Task I: Lipid profile leading to the first HF diagnosis:* Figure 3 presents the SHAP beeswarm plots from prediction task I for male and female patients separately, at the 12-, 24-, and 36-month time points. In beeswarm plots, lipid biomarkers are presented from top to bottom according to their mean absolute SHAP values. For male patients, LDL-C and ApoB are invariably the two most influential biomarkers that predict the outcomes of HF, followed by HDL-C and TG. The Shapley values for LDL-C from patient records are color coded to show that high values in red fall on the negative side of the outcome, whereas low values in blue are on the positive side. LDL-C has the strongest influence among the four lipid biomarkers, indicating that lower values predict a higher likelihood of developing HF in the future, confirming the paradoxical observation about LDL-C values in the literature. Unlike LDL-C, the Shapley values for ApoB show mixed directions, with low values extending in both directions of the HF outcome. In the 36-month prediction model, elevated ApoB is associated with a higher likelihood of HF events, but this effect diminishes as the time approaches the first HF diagnosis. Low HDL-C levels strongly predict HF outcomes within a 12-month window, but this association becomes less clear at longer time windows. At the 36-month prediction window, a higher TG level is associated with an increased risk of a future HF event, but this relationship reverses when the prediction window is shortened to 12 months. It is worth noting that the predictive performance of task-I is not particularly strong, likely because HF records can fall into either the positive or negative group depending on the selected time window. In female patients, the inverse relationship between LDL-C levels and HF outcomes (also known as the cholesterol paradox) is more prominent than in their male counterparts, especially in the 24- and 36-month prediction windows. The association between ApoB and the first HF event is equally ambiguous in the female cohort, except that low ApoB levels strongly predict a lower likelihood of HF at the 12-month window. Unlike their male counterparts, HDL and TG show unambiguous trends across the three time windows. In clinical practice, it is well established that elevated HDL and low triglyceride levels are associated with a reduced risk of a first heart failure event [25].
2. *Task II: Predicting the first HF diagnosis against the non-CVD cohort:* Figure 4 shows the beeswarm plot for prediction task II. The smooth values of LDL-C show a consistent trend in all time windows, regardless of biological sex. The median levels of LDL-C in purple show the weakest association with the prediction of HF, while very high levels in red cancel out the predictive value in both directions. Consistent with the cholesterol paradox, lower LDL-C levels (blue) are again associated with a higher likelihood of predicting HF in both men and women. The pattern of ApoB is somewhat unclear in both male and female cohorts; however, higher levels of ApoB generally correspond to a higher probability of HF diagnoses, and vice versa.

**Fig. 3.**
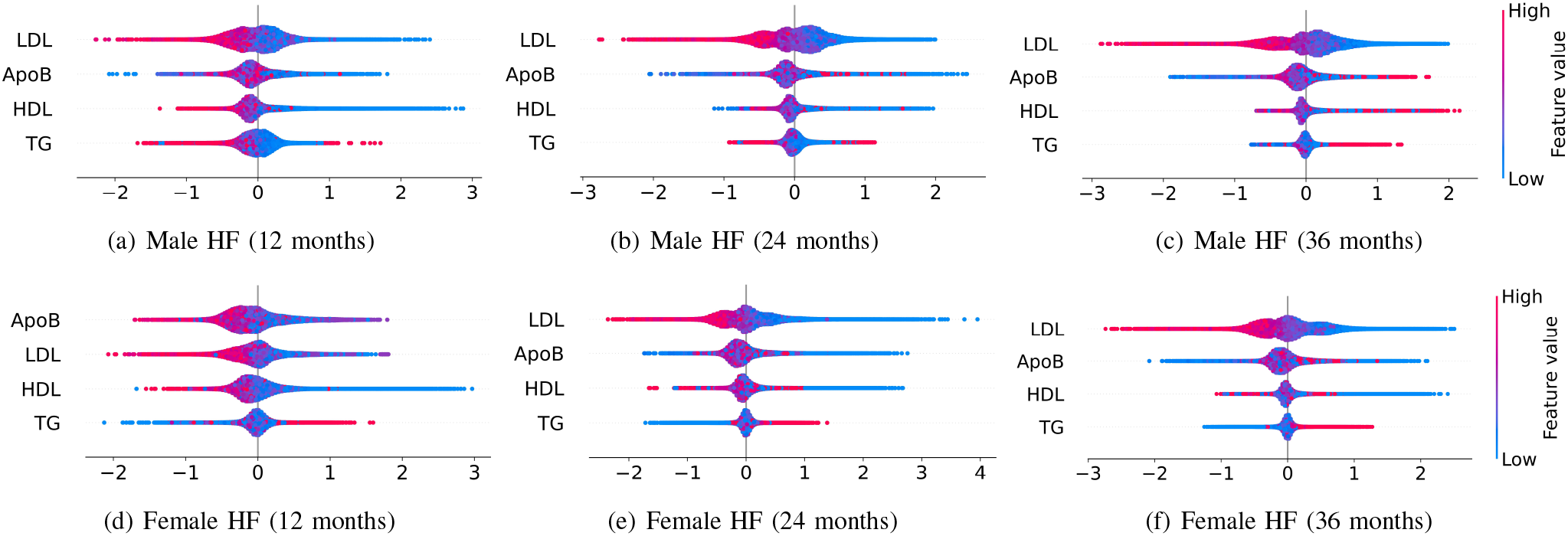
SHAP beeswarm plots from prediction task I, predicting the first heart failure event at varying time windows.

**Fig. 4.**
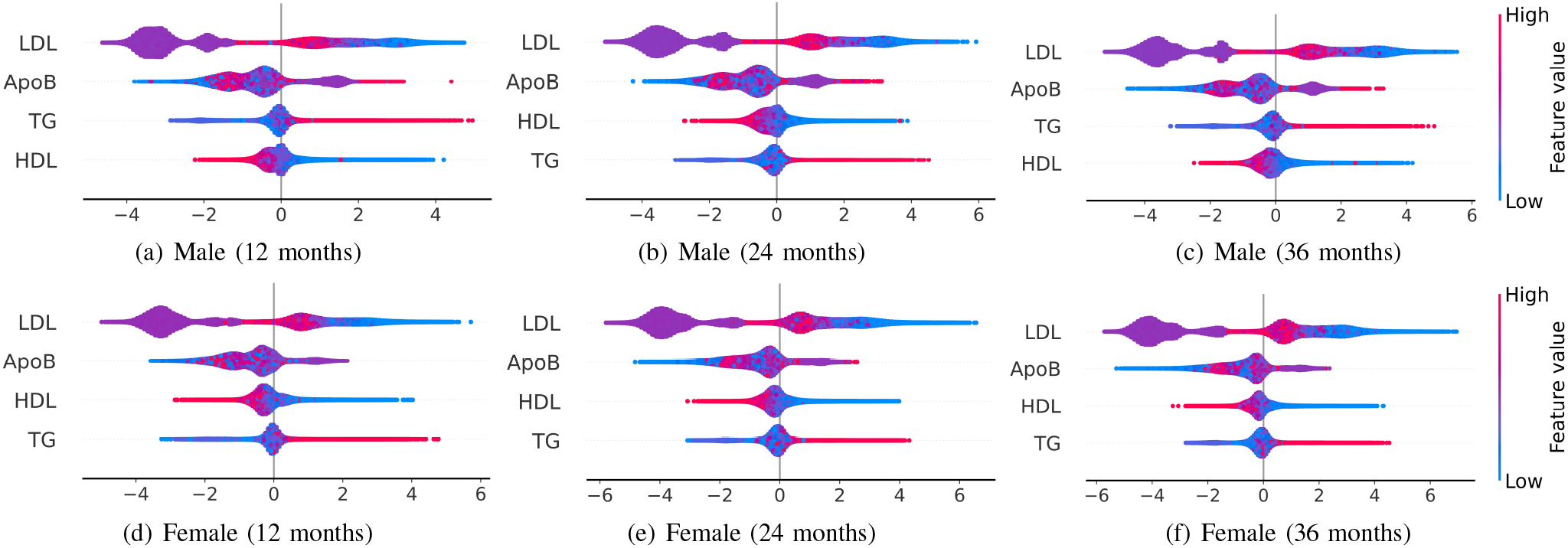
SHAP beeswarm plots from prediction task II, predicting the first heart failure event against patients with no records of cardiovascular diseases at varying time windows.

Unlike Task I, TG and HDL are comparable in influence and alternate as the third- and fourth-ranked biomarkers, with TG ranking above HDL in male patients at the 12- and 36-month windows. When analyzed separately, both male and female groups show clear, robust trends for TG and HDL, unlike Task I. High TG levels are consistently linked to an increased probability of HF diagnosis across all time windows, whereas low TG levels are associated with a reduced likelihood. In contrast, low HDL is clearly associated with a higher likelihood of HF, while elevated HDL levels are generally associated with a reduced risk of developing CVD, including HF. These observations about HDL-C and TG align with the notion that metabolic parameters play a critical role in cardiovascular disease.

## V. Discussion

This study investigates how lipid biomarkers, including estimated ApoB, predict first heart failure events across six pre-diagnosis windows. The findings of the paper can be summarized as follows.

First, lipid biomarkers behave differently across two separate tasks. They provide only a modest signal in the prospective HF cohort (Task I, combined AUROC 0.677 to 0.627), but show a far clearer separation when HF patients are compared with a non-CVD population (Task II, AUROC 0.919–0.937 in sex-stratified cohorts). Second, population-level stratification clearly illustrates the cholesterol paradox: patients with HF have lower—not higher—LDL-C, HDL-C, and estimated ApoB than patients without CVD. Third, elevated TG and reduced HDL levels, reflecting poor metabolic health, consistently predict initial HF events. Fourth, lipid signaling is highly sex-dependent, and sex-separated analyses markedly improve population-level discrimination compared to the pooled cohort (0.919–0.937 vs. 0.728–0.762).

### A. Task I versus Task II

In Task I, lipid profiles are not informative in prospective HF cohorts to predict the first HF event. Low precision and recall, with higher specificity, indicate many false negatives. The high number of false negatives suggests that in this cohort, lipid profiles are not effective in prospectively identifying individuals who would develop HF. This may be due to how the prediction windows are set up, where the negative class may still include patients diagnosed with HF later. The combination of low F1 scores and moderate ROC scores, even with limited class imbalance, suggests that the F1 threshold likely needs adjustment to improve performance. Comparable AUROC and accuracy values further suggest minimal class imbalance.

Task II strictly separates patients with prospective HF diagnosis and those who would not receive any CVD diagnosis in the future. This clear distinction may enable lipid profiles to serve as strong predictors, as evidenced by their consistently high prediction performance scores. A relatively low precision against a higher recall indicates a large number of false positives. The high number of false positives suggests that the non-CVD cohort carries a lipid profile that is indistinguishable from that of patients who develop HF. Accordingly, precision and F1 scores improve steadily as the pre-DW lengthens from 6 to 36 months, following the increasing prevalence of positive records. Additionally, recall and AUROC remain high and stable in all time windows. This stability indicates that the lipid signal that separates prospective HF patients from the non-CVD population does not depend on the choice of the prediction window, unlike task I.

## VI. Conclusion

This paper presents a multi-window, two-task framework to predict first heart failure events from lipid panels. Using EHR data from the All of Us Research Program, it combines estimated ApoB and standard biomarkers in an explainable XGBoost model. Lipids are only modestly discriminative within a prospective heart failure cohort, but they separate heart failure from a non-CVD population much better after sex stratification. The population-level comparison illustrates the cholesterol paradox recovered by the model: lower LDL-C shifts predictions toward heart failure years before onset, while estimated ApoB adds complementary information. Because lipid signals are sex-dependent, sex-specific models outperform pooled cohorts. Overall, lipid-based heart failure risk assessment is most informative when multi-biomarker, sexspecific, and temporally aware.

## Data Availability

This study used secondary, de-identified human-subject data available through the All of Us Research Program's credentialed repository in the United States.

https://support.researchallofus.org/hc/en-us/articles/360039175232-Intro-to-the-All-of-Us-Researcher-Workbench

## Acknowledgment

We gratefully acknowledge All of Us participants for their contributions to the All of Us health data repository. These real-world medical data are invaluable in the advancement of the study of AI and statistical models. We also thank the National Institutes of Health’s All of Us Research Program for making available the participant data examined in this study.

## Ethics and Consent to Participate

The manuscript uses secondary and de-identified human subject data recorded in the NIH-sponsored All of Us Research Program. No human subjects were recruited for the investigation. Ethics and consent to participation are not applicable.

## Author Contribution declaration

Yina Hou contributed to Data curation, Formal Analysis, Validation, Visualization, Writing - Original draft preparation, and Investigation. Md Kamrozzaman Bhuiyan contributed to Formal Analysis, Validation, and Writing – Review and Editing. Manar D. Samad contributed to Conceptualization, Methodology, Funding acquisition, Supervision, Writing – Review and Editing, Investigation, Project administration

